# Metabolic Dysfunction-Associated Fibrosis Score (MAF-5) and Gallstone Prevalence: A Cross-Sectional Analysis Using NHANES 2017–2020 Data

**DOI:** 10.64898/2026.08.02.26359499

**Authors:** Yujian Huang, Junlin Lu, Hao Wang

**Author notes:** **Correspondence:** Hao Wang, College of Animal Science and Veterinary Medicine, Henan Institute of Science and Technology, Xinxiang, Henan, China.

## Abstract

**Background:** Gallstones are one of the most common gastrointestinal conditions closely associated with metabolic dysfunction. The metabolic dysfunction-associated fibrosis-5 (MAF-5) score has been established as a non-invasive indicator for assessing liver fibrosis in individuals with metabolic abnormalities. However, comprehensive large-scale research examining the correlation between the MAF-5 score and gallstone occurrence remains limited. This study seeks to clarify the link between the MAF-5 score and gallstone prevalence using nationally representative data from the National Health and Nutrition Examination Survey (NHANES).

**Methods:** This study examined data from 15,560 NHANES 2017–2020 participants aged 20 years or older, ensuring complete records for MAF-5 scores and gallstone status. Gallstone presence was identified through self-reported physician diagnoses. To assess the relationship between MAF-5 scores and gallstone prevalence, weighted logistic regression models were applied, adjusting for demographic characteristics, lifestyle factors, and health conditions. Subgroup analyses were conducted to evaluate the stability of this association and detect possible interactions. Sensitivity analyses were performed by excluding extreme values (±3SD) to assess result robustness. Furthermore, MAF-5 scores were divided into quartiles to investigate gallstone prevalence trends, and a restricted cubic spline (RCS) model was employed to visualize response patterns.

**Results:** A total of 15,560 participants met the inclusion criteria, with 747 in the gallstone group and 6,367 in the non-gallstone group. MAF-5 scores were significantly higher in the gallstone group (P < 0.001). After adjusting for multiple covariates, each unit increase in MAF-5 score correlated with a 14% higher gallstone prevalence (OR = 1.14, 95% CI: 1.06–1.23). Quartile-based analysis indicated that individuals in the highest MAF-5 quartile had a 2.12-fold higher prevalence of gallstones than those in the lowest quartile (OR = 2.12, 95% CI: 1.13–3.98). RCS analysis confirmed a linear association between MAF-5 scores and gallstone prevalence. Subgroup analyses showed this association remained stable across age, sex, and racial/ethnic groups, with no significant interactions. Sensitivity analyses, excluding extreme values (±3SD), reinforced the reliability of these findings (OR = 1.15, 95% CI: 1.04–1.28).

**Conclusion:** The MAF-5 score is significantly and positively associated with gallstone prevalence, independent of demographic and lifestyle confounders. These findings indicate that the MAF-5 score may be a useful tool for assessing gallstone prevalence in individuals with metabolic dysfunction, offering valuable insights for early screening and targeted health management strategies.

## 1. Introduction

Cholelithiasis, the formation of gallstones within the gallbladder or bile ducts, is among the most common biliary disorders worldwide[1]. Epidemiological studies estimate that gallstones affect approximately 10% to 15% of the global population[2], with prevalence rising due to aging, obesity, and the increasing burden of metabolic dysfunction[3, 4].In China, gallstone prevalence varies significantly across regions, with high-risk populations—particularly individuals with obesity and diabetes— exhibiting a markedly higher prevalence than the general population[5, 6]. Beyond biliary colic and digestive disturbances, gallstones are strongly associated with complications such as cholecystitis, cholangitis, pancreatitis, and gallbladder cancer[7], significantly reducing quality of life and imposing substantial healthcare and socioeconomic burdens. Given the chronic nature of gallstone disease and its recurrence risk, effective prevention and management strategies are critical. Nurses play a key role in patient education, particularly in guiding dietary modifications and lifestyle adjustments to reduce gallstone formation and postoperative complications. Studies suggest that nurse-led interventions improve patient adherence to dietary recommendations and enhance long-term metabolic health, which is essential for gallstone prevention[8, 9]. However, despite advances in clinical management, the identification of high-risk individuals remains challenging, necessitating the development of improved screening tools.

Metabolic dysfunction is a well-recognized contributor to gallstone formation, with conditions such as obesity, insulin resistance, and type 2 diabetes significantly elevating gallstone risk[10, 11]. These metabolic disturbances interfere with hepatic cholesterol and bile acid metabolism while also impairing gallbladder motility, ultimately promoting bile stasis and gallstone development[12]. Furthermore, metabolic dysfunction is frequently linked to chronic low-grade inflammation and liver fibrosis, both of which play key roles in bile acid dysregulation—an essential mechanism in gallstone pathogenesis[13]. This evidence underscores the involvement of the metabolic-liver-gallbladder axis in gallstone formation. In 2024, the Metabolic Dysfunction-Associated Fibrosis Score (MAF-5) was introduced as a non-invasive measure for evaluating liver fibrosis in individuals with metabolic dysfunction[14]. Compared with conventional fibrosis scores such as FIB-4 and NFS, MAF-5 more accurately reflects metabolic-associated liver disease (MASLD) pathology, making it particularly suitable for metabolically abnormal populations[15]. Studies have demonstrated that MAF-5 effectively predicts liver fibrosis in MASLD patients, including younger individuals with metabolic dysfunction[16]. Although growing evidence suggests a link between liver fibrosis and gallstone formation—potentially through disruptions in bile acid metabolism and gallbladder motility—the utility of MAF-5 as a screening tool for gallstone prevalence remains largely unexplored.

To bridge this gap, the present study leverages nationally representative data from NHANES 2017–2020 to systematically investigate the association between MAF-5 scores and gallstone prevalence. This study aims to provide robust epidemiological evidence on the interplay between metabolic dysfunction, liver fibrosis, and gallstone disease while also offering novel insights into early gallstone risk identification in metabolically abnormal populations.

## 2. Methods

### 2.1 Survey Description

This study utilized NHANES data, a cross-sectional survey designed to provide a representative evaluation of the U.S. population’s health and nutrition. NHANES employs a multistage, stratified sampling methodology to ensure national representativeness. Conducted by the National Center for Health Statistics (NCHS), the survey was approved by the Institutional Review Board (IRB), with all participants providing written informed consent in compliance with ethical guidelines.

### 2.2 Study Population

The study cohort was derived from NHANES 2017–2020 survey cycles. Participants were eligible if they met the following criteria: aged ≥20 years, availability of complete MAF-5 score data, and full documentation of gallstone diagnosis. Only individuals fulfilling all inclusion requirements were retained for the final analytical dataset.

### 2.3 Definition of MAF-5 Score

The Metabolic Dysfunction-Associated Fibrosis Score (MAF-5) is a non-invasive tool designed to assess fibrosis in individuals with metabolic dysfunction. Developed based on prior research, this scoring system incorporates multiple metabolic parameters, including body mass index (BMI), waist circumference (WC), the BMI-WC interaction, diabetes history (yes/no), the natural logarithm of aspartate aminotransferase (AST), and platelet count. The MAF-5 score is calculated using the following formula:

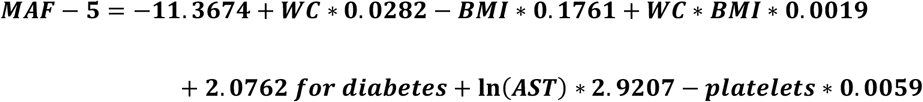

where WC is measured in centimeters (cm), BMI in kg/m², AST in U/L, and platelets in 109/L10^9/L109/L. Diabetes is a binary variable (0 = No, 1 = Yes), and ln denotes the natural logarithm function.

### 2.4 Definition of Gallstones

Gallstone status in this study was ascertained using self-reported data from the NHANES questionnaire. Specifically, participants responded to the question: “Has a doctor ever informed you that you have gallstones?” Participants who responded “Yes” were assigned to the gallstone group, while those who answered “No” were designated as the non-gallstone group. The self-reported gallstone diagnosis method has been extensively used in large-scale epidemiological studies and has demonstrated high reliability[17, 18].

### 2.5 Covariates

To account for potential confounders, this study included demographic, lifestyle, and health status variables in the analytical models. Demographic variables comprised age, sex, race/ethnicity, and educational level. Lifestyle factors included smoking status, alcohol consumption, and physical activity levels. Smoking status was determined using NHANES questionnaire data, with individuals reporting a lifetime consumption of at least 100 cigarettes classified as smokers. Alcohol consumption was evaluated based on self-reported intake frequency in the past year, with non-drinkers defined as those who reported abstinence. Physical activity levels were assessed using the Global Physical Activity Questionnaire (GPAQ), with metabolic equivalent task (MET, min/week) computed as the product of the activity intensity coefficient (WET), weekly frequency, and session duration. Participants with MET values below 600 min/week were categorized as physically inactive. Health status variables encompassed hypertension and lipid profile markers. Hypertension was defined as either a documented physician diagnosis in NHANES or self-reported hypertension. Lipid profile markers, including high-density lipoprotein cholesterol (HDL-C) and triglycerides (TG), were derived from laboratory measurements.

### 2.6 Statistical Analysis

A cross-sectional analysis was performed using data from the NHANES 2017–2020 survey cycle. Participants were categorized into two groups based on gallstone status, and their demographic characteristics, lifestyle factors, and health-related attributes were analyzed. Continuous variables were presented as mean ± standard deviation (SD), while categorical variables were reported as frequencies and percentages (%). The association between MAF-5 scores and gallstone prevalence was assessed using both univariate and multivariate logistic regression models, with results expressed as odds ratios (ORs) and 95% confidence intervals (CIs). To examine trends in gallstone prevalence across MAF-5 levels, participants were stratified into quartiles (Q1–Q4), and prevalence rates were compared across quartile groups. Additionally, restricted cubic spline (RCS) modeling was employed to explore potential nonlinear relationships between MAF-5 scores and gallstone prevalence. Subgroup analyses were conducted to evaluate whether this association remained consistent across different age, sex, and BMI subgroups, while sensitivity analyses were performed by excluding extreme MAF-5 values (±3SD) to assess the robustness of the findings. All statistical analyses were conducted using R software (version 4.2.3), with a two-tailed P-value <0.05 considered statistically significant.

## 3. Results

### 3.1 Baseline Characteristics

A total of 15,560 participants satisfied the inclusion criteria, among whom 747 had a confirmed diagnosis of gallstones, while 6,367 had no reported history of the condition. Among these, 7,114 had complete data for all covariates and were included in the multivariable models. The sample selection process and baseline characteristics are outlined as follows (Figure 1, Table 1). Comparative analysis revealed that individuals in the gallstone group tended to be older and included a higher proportion of females and non-Hispanic White participants relative to the non-gallstone group. In terms of lifestyle behaviors, smoking and alcohol consumption were more frequently observed in the gallstone group than in the non-gallstone group. Furthermore, the MAF-5 score was markedly higher in the gallstone group, implying a possible association between metabolic dysfunction and gallstone formation.

**Figure 1.**
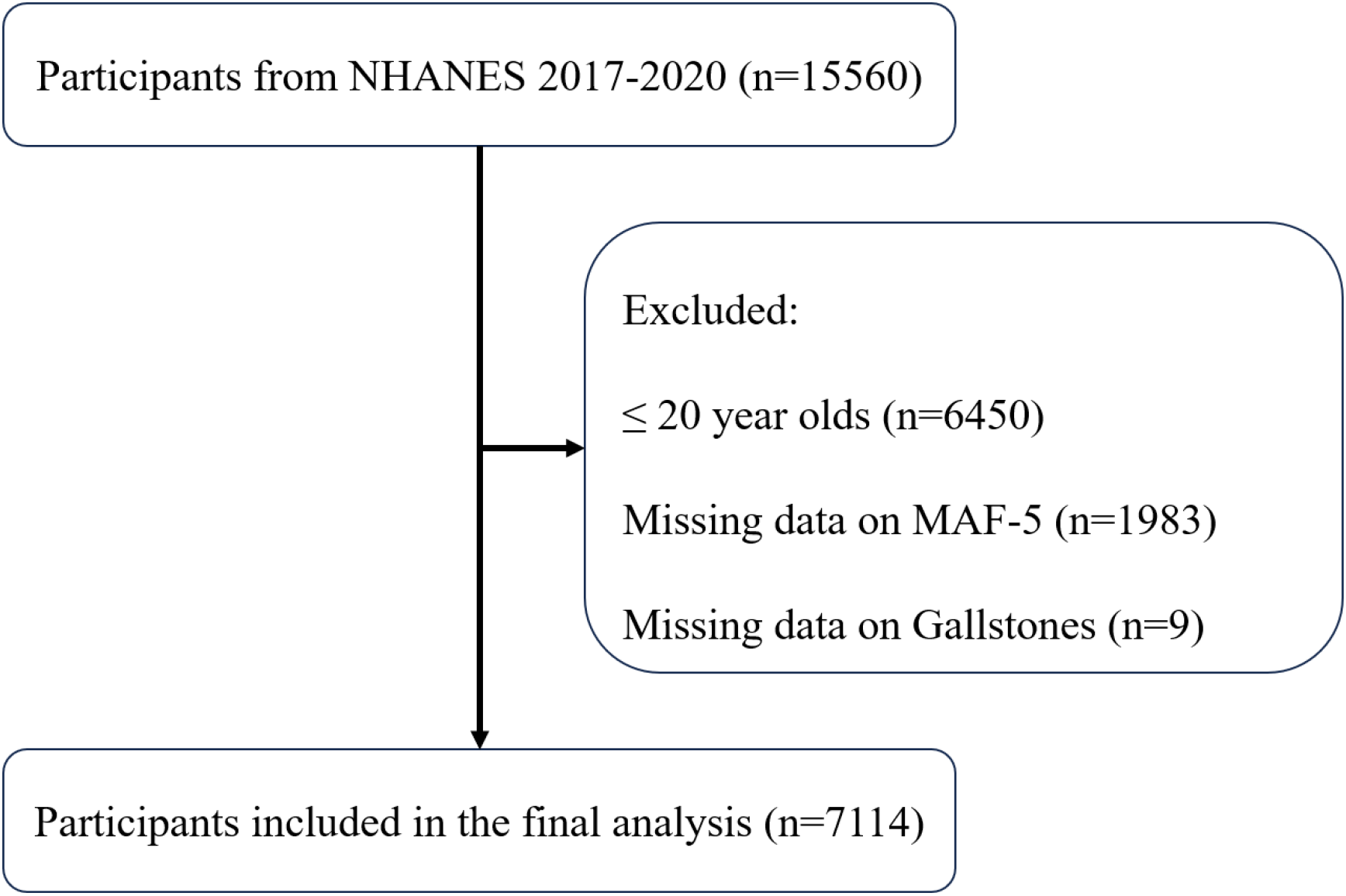
Flowchart of Participant Inclusion Process.

**Table 1.**
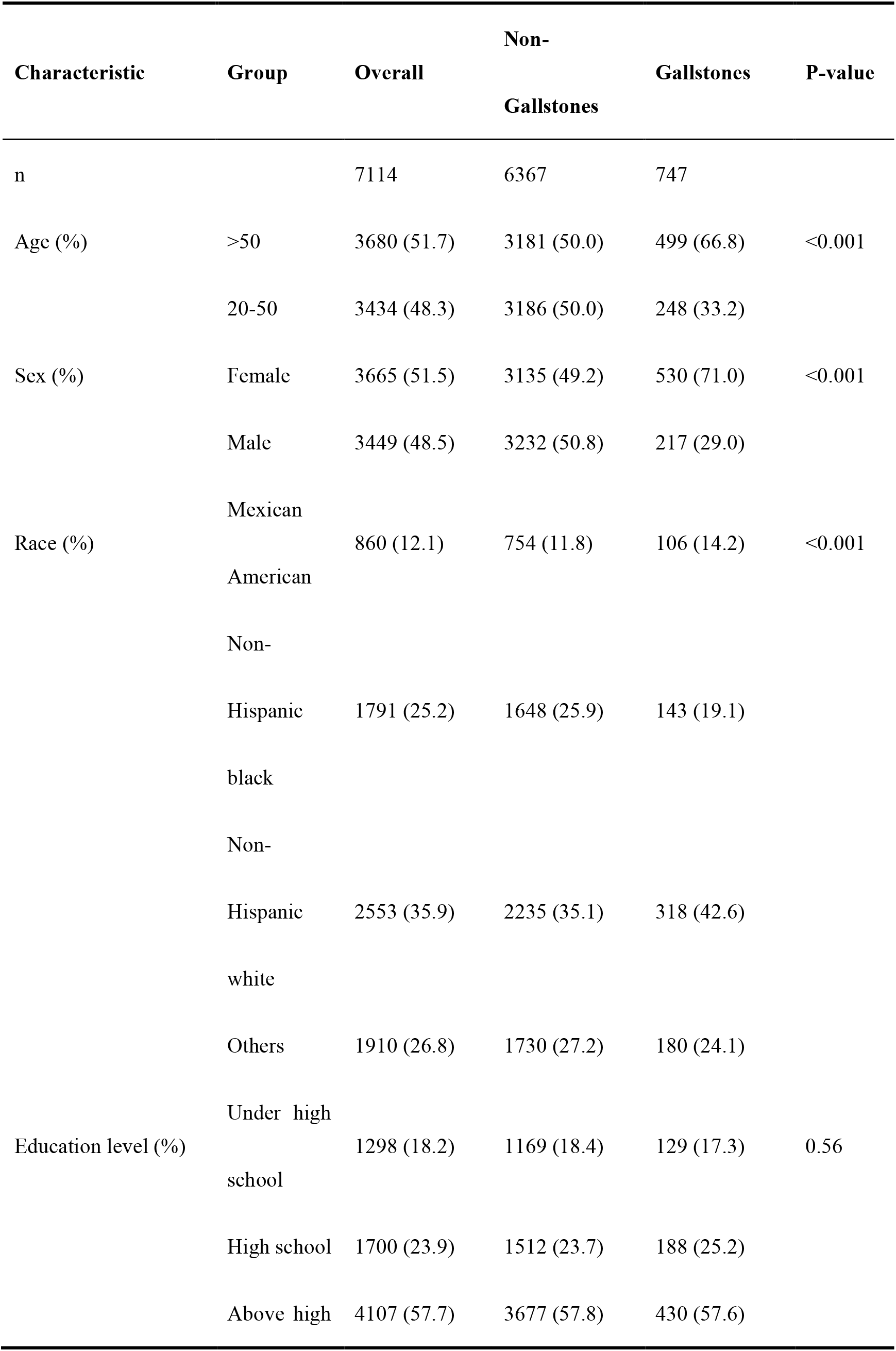
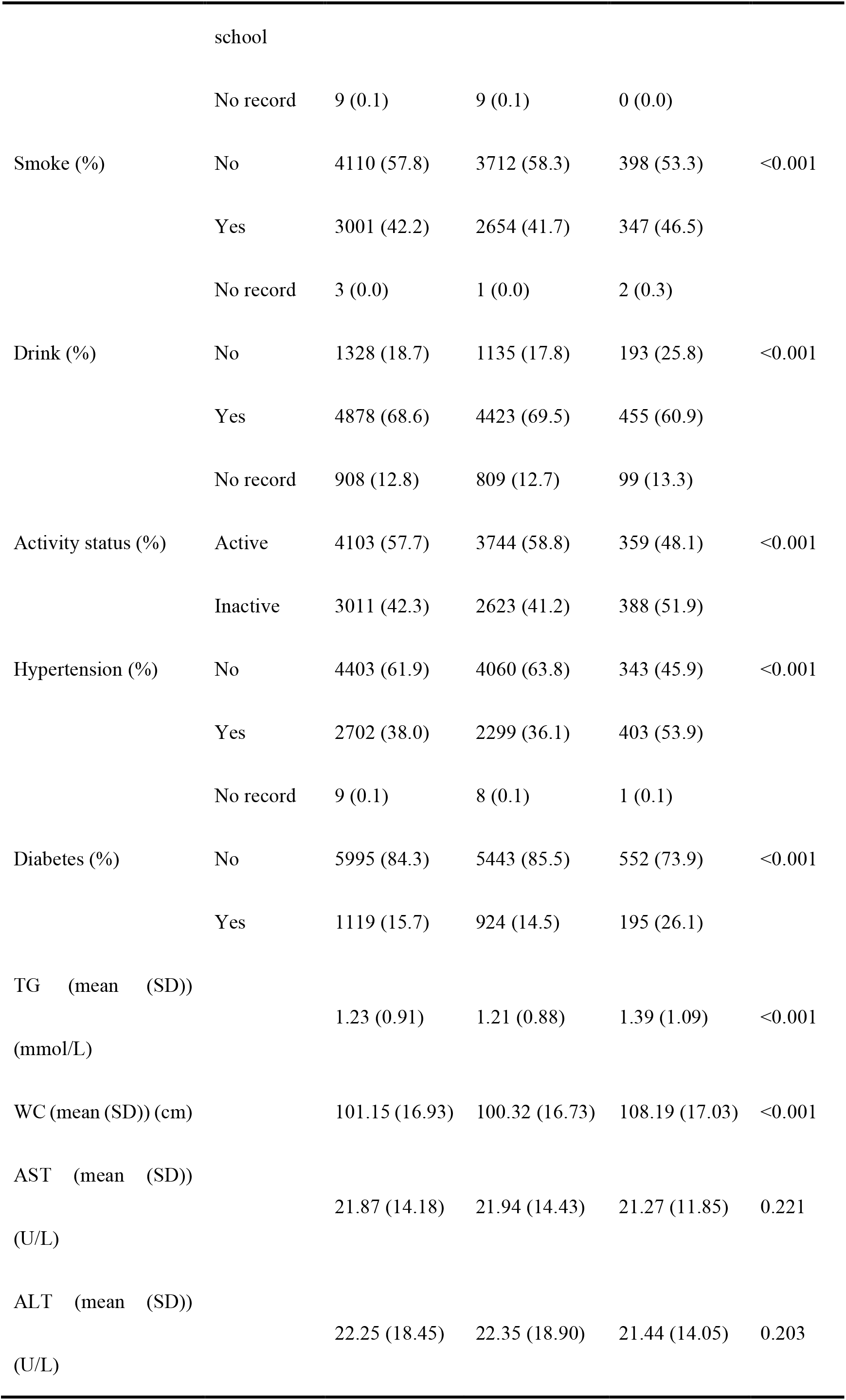
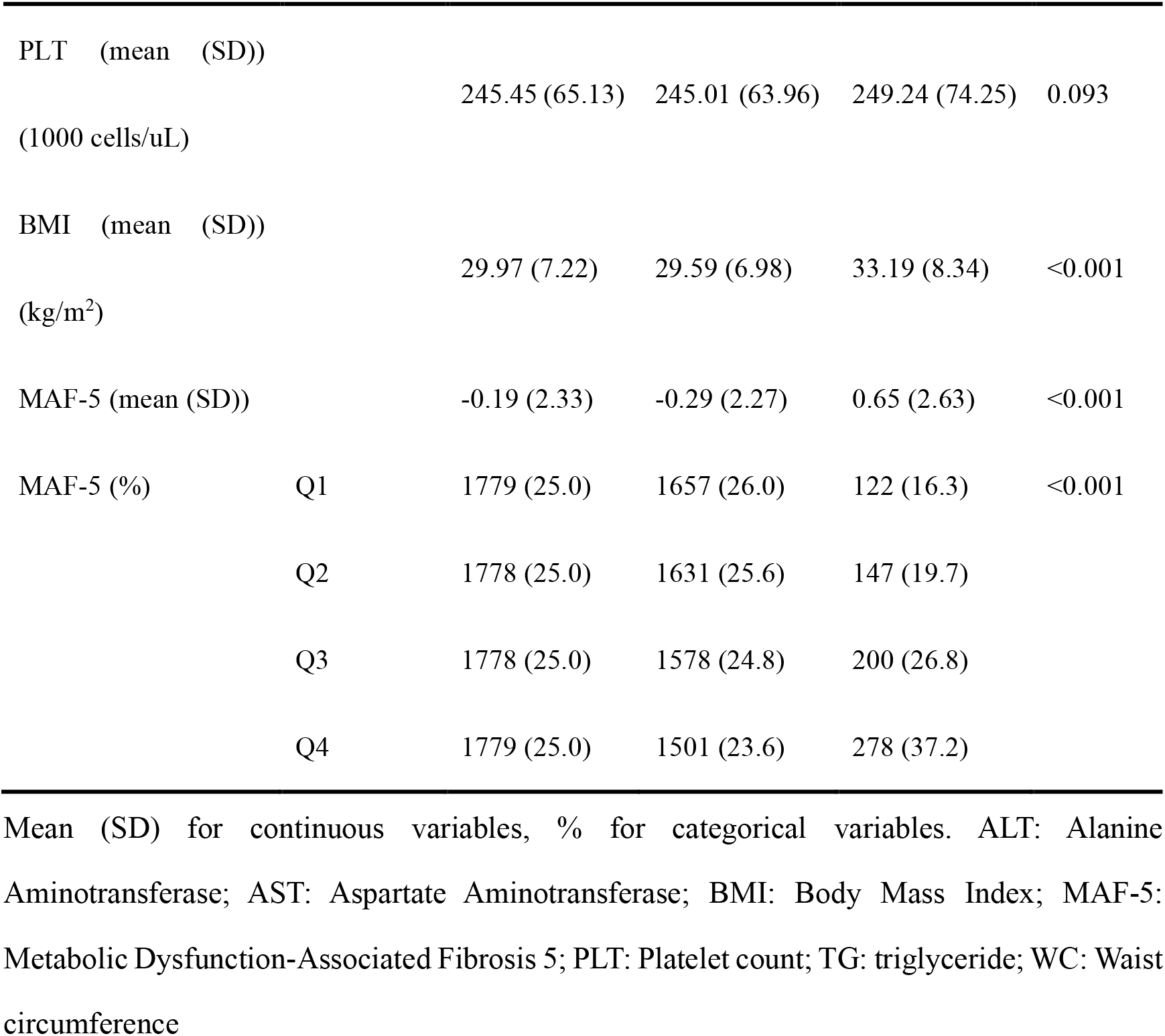
Baseline characteristics of the study population.

### 3.2 Association Between MAF-5 Score and Gallstone Prevalence

Logistic regression models were utilized to examine the relationship between MAF-5 scores and gallstone prevalence (Table 2). In the unadjusted analysis, each one-unit increase in MAF-5 score was associated with a 15% higher prevalence of gallstones (OR = 1.15, 95% CI: 1.10–1.20). After adjusting for demographic characteristics, lifestyle behaviors, and health conditions in the fully adjusted model (Model 3), this association remained significant, with each unit increment in MAF-5 score corresponding to a 14% increase in gallstone prevalence (OR = 1.14, 95% CI: 1.06–1.23), reinforcing its independent correlation with gallstone occurrence. Quartile-based analysis indicated that individuals in the highest MAF-5 quartile (Q4) had more than double the prevalence of gallstones compared to those in the lowest quartile (Q1) (OR = 2.12, 95% CI: 1.13–3.98). Additionally, restricted cubic spline (RCS) modeling confirmed a linear positive relationship between MAF-5 scores and gallstone prevalence, illustrating a continuous upward trend as MAF-5 scores increased (Figure 2).

**Figure 2.**
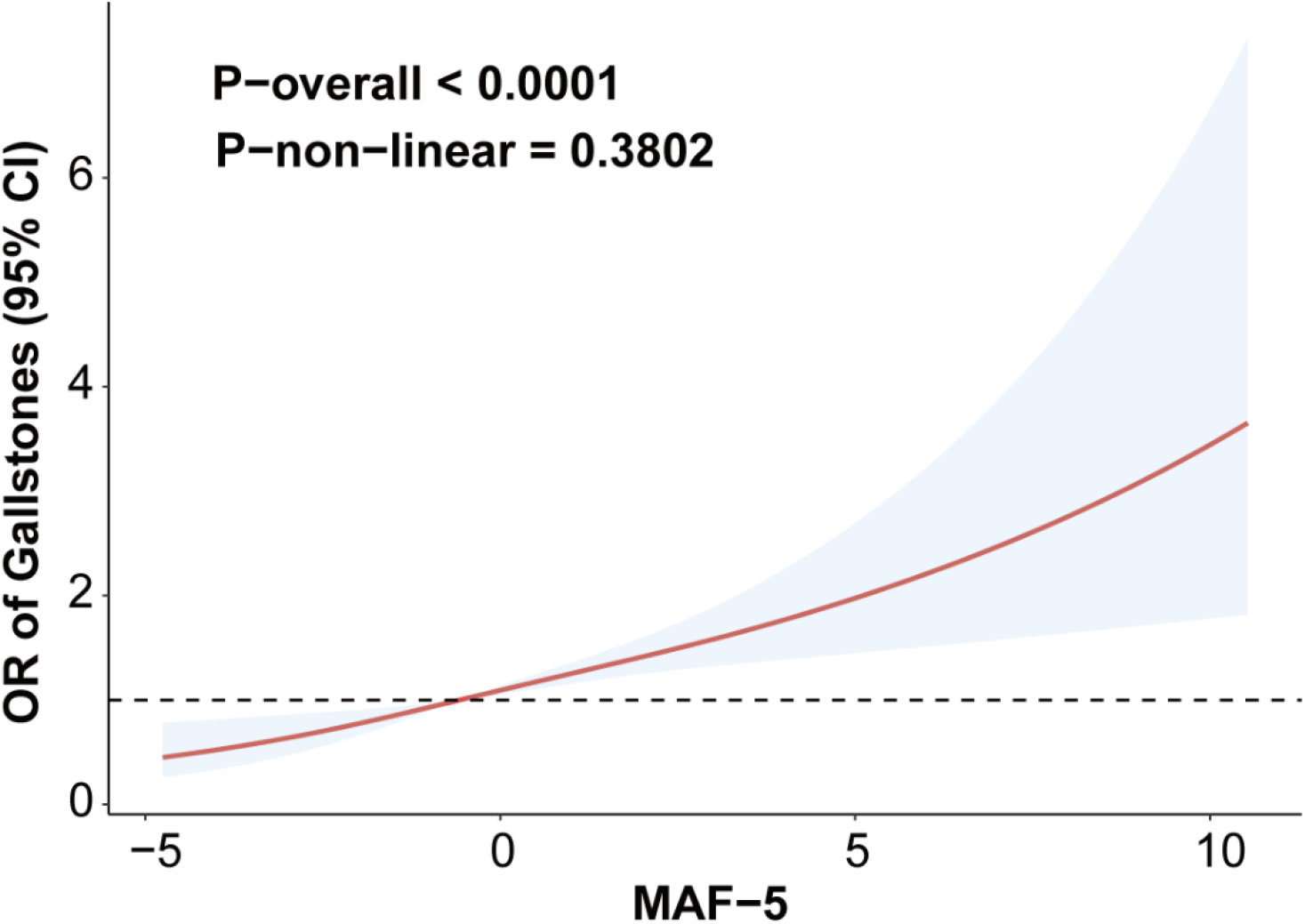
RCS curve depicting the association between MAF-5 Score and gallstone prevalence.

**Table 2.**
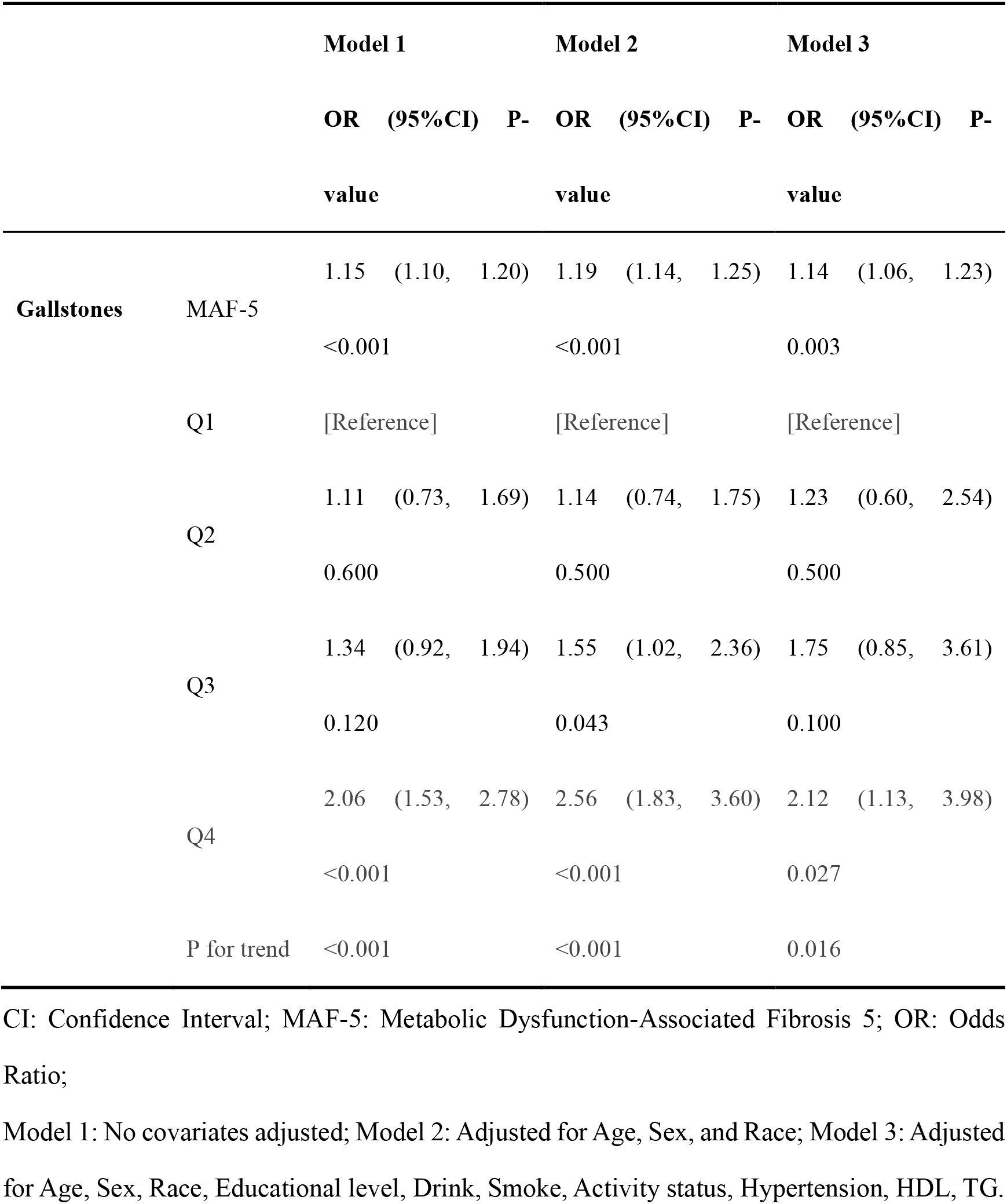
Association Between MAF-5 Score and Gallstone Prevalence.

### 3.3 Subgroup Analysis

Stratified analyses were performed across subgroups defined by sex, age, and race/ethnicity (Table 3). The results demonstrated that the positive association between MAF-5 scores and gallstone prevalence remained consistent across all subgroups, with no statistically significant heterogeneity detected. Moreover, interaction analysis revealed no statistically significant interactions, suggesting that this association is stable and generalizable across diverse populations.

**Table 3.** Subgroup analysis of the association between MAF-5 score and gallstone prevalence.

| Characteristic | Group | OR (95%CI) P-value | P for interaction |
| --- | --- | --- | --- |
| Age | 20-50 | 1.11 (1.02, 1.22) 0.022 | 0.6 |
|  | >50 | 1.17 (1.05, 1.32) 0.010 |  |
| Sex | Male | 1.19 (1.05, 1.36) 0.012 | 0.6 |
|  | Female | 1.12 (1.03, 1.22) 0.016 |  |
| Race | Mexican American | 1.00 (0.77, 1.29) >0.900 | 0.2 |
|  | Non-Hispanic black | 1.19 (1.08, 1.31) 0.003 |  |
|  | Non-Hispanic white | 1.14 (1.02, 1.28) 0.027 |  |
|  | Others | 1.20 (1.03, 1.40) 0.024 |  |
| Smoke | No | 1.07 (0.95, 1.21) 0.200 | 0.2 |
|  | Yes | 1.22 (1.13, 1.32) <0.001 |  |
| Drink | No | 1.23 (1.08, 1.40) 0.005 | 0.2 |
|  | Yes | 1.14 (1.05, 1.23) 0.004 |  |
| Hypertension | No | 1.12 (1.01, 1.25) 0.039 | 0.6 |
|  | Yes | 1.17 (1.05, 1.31) 0.011 |  |

CI: Confidence Interval; OR: Odds Ratio

**Table 4.**
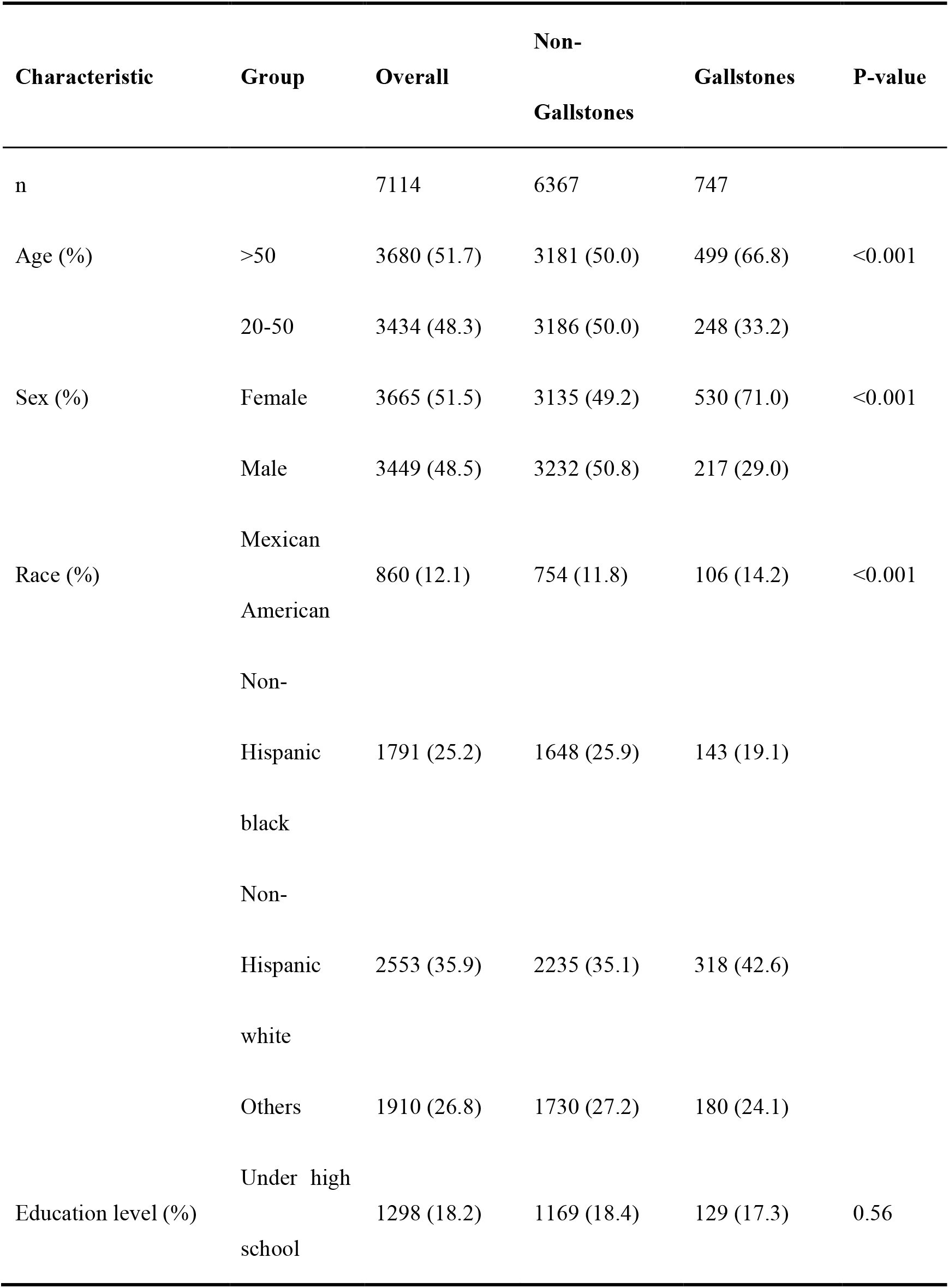
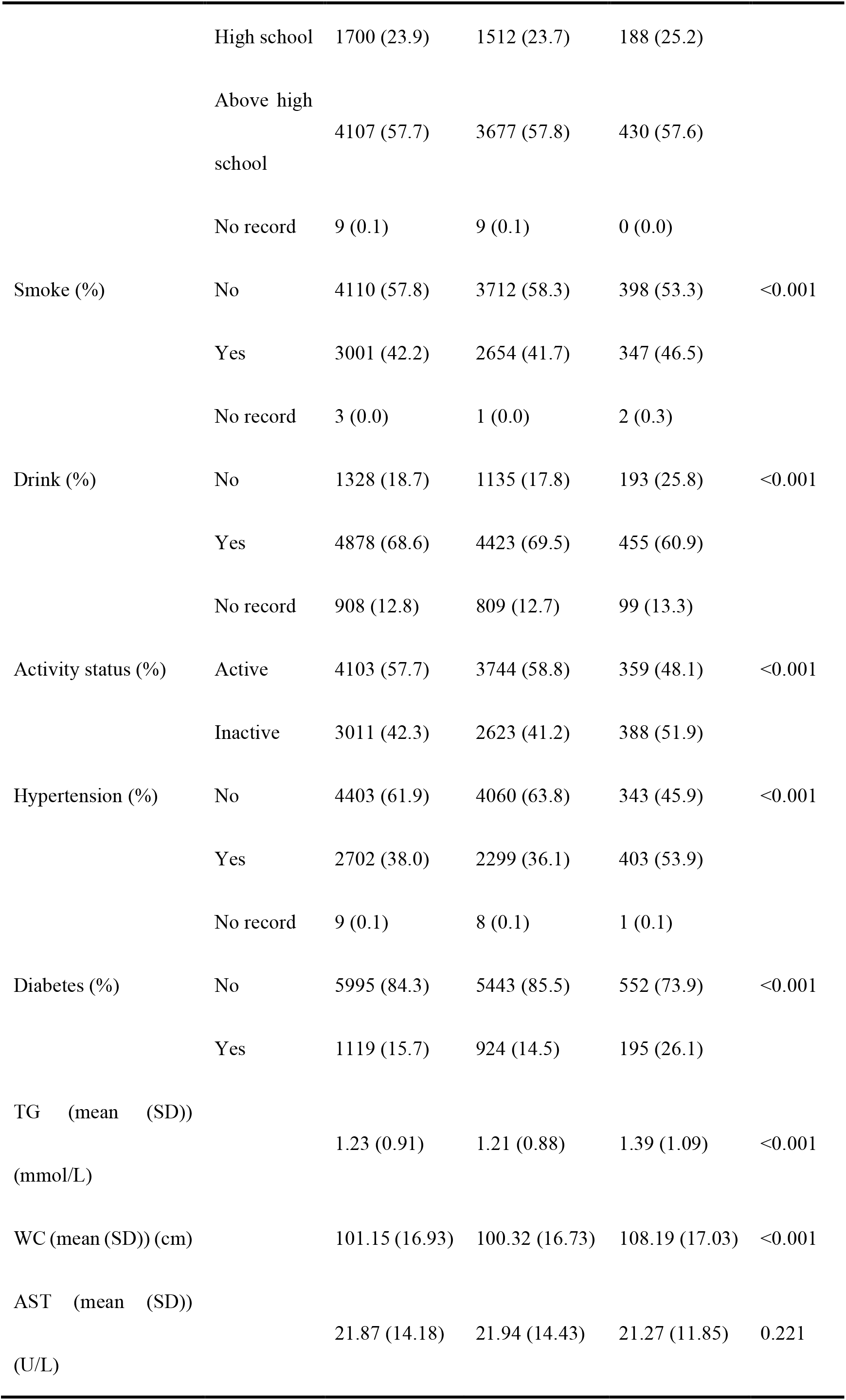
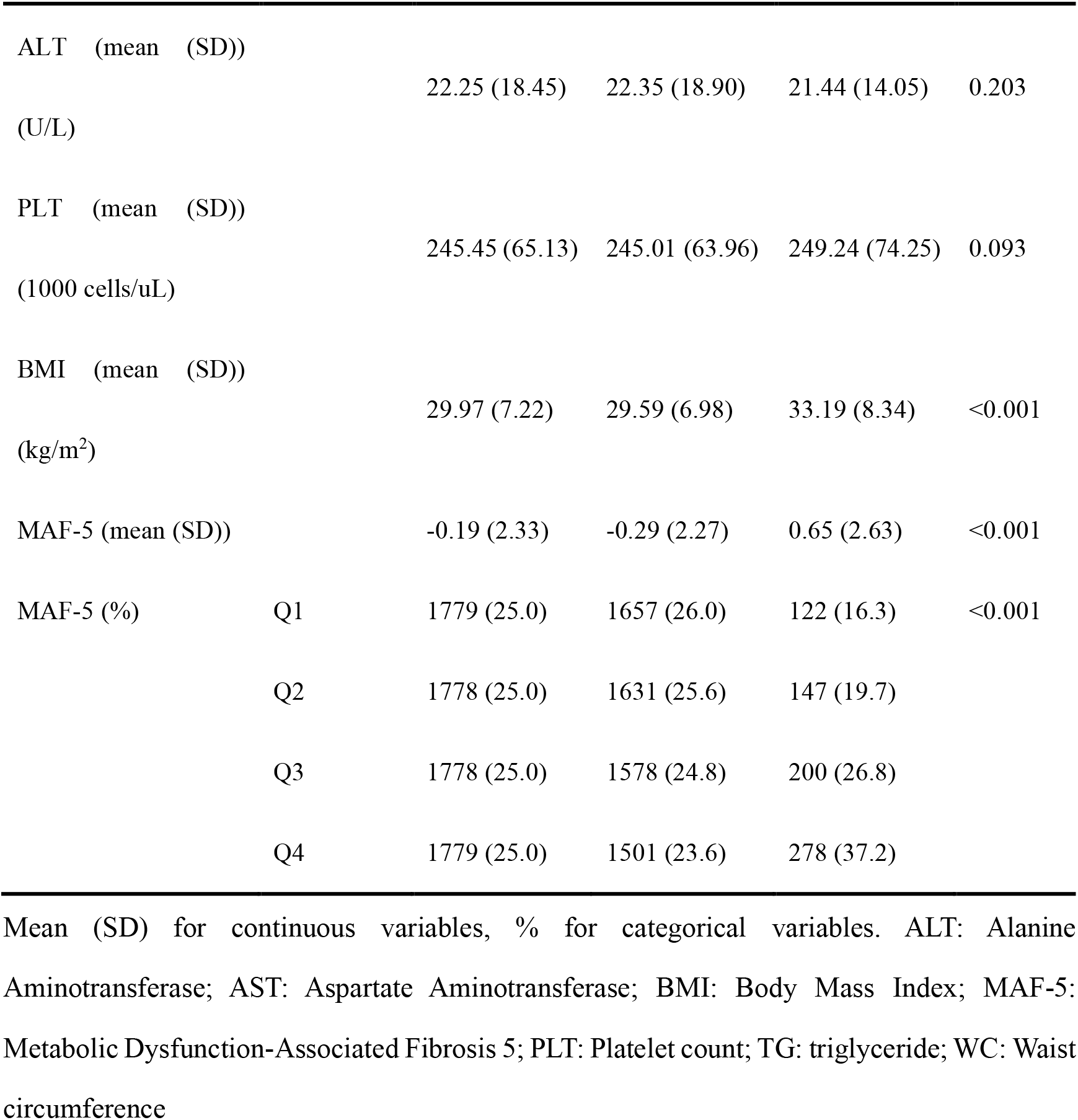
Baseline characteristics of the study population.

### 3.4 Sensitivity Analysis

To assess the robustness of the findings, an additional sensitivity analysis was conducted by excluding extreme MAF-5 values (±3SD), yielding a refined dataset of 7,037 participants (including 730 in the gallstone group and 6,307 in the non-gallstone group). Following reanalysis, the association between MAF-5 scores and gallstone prevalence remained statistically significant. Each additional unit increase in MAF-5 score was associated with a 15% higher gallstone prevalence (OR = 1.15, 95% CI: 1.04–1.28) in the fully adjusted model. These results were consistent with the primary analysis (Supplementary Table 1), further reinforcing the reliability and coherence of the findings.

## 4. Discussion

This study leveraged NHANES 2017–2020 data to comprehensively examine the link between MAF-5 scores and gallstone prevalence. The results demonstrated a strong correlation between elevated MAF-5 scores and a higher prevalence of gallstones, which persisted even after adjusting for multiple confounders. Subgroup and sensitivity analyses further validated the consistency of this relationship across different age groups, sexes, and racial/ethnic backgrounds, emphasizing its stability and applicability. These findings indicate that the MAF-5 score may function not only as a marker for metabolic dysfunction-associated liver fibrosis but also as a valuable screening tool for identifying individuals at an increased prevalence of gallstones. For those with metabolic dysfunction and elevated MAF-5 scores, early detection and comprehensive gallbladder health management should be prioritized to reduce the likelihood of gallstone development.

Previous studies have extensively documented the strong association between metabolic dysfunction and gallstone formation. Obesity, a well-established risk factor for gallstones, has been validated in multiple prospective cohort studies. Research suggests that each 5 kg/m² increment in BMI is associated with a 20%–30% increase in gallstone risk[1]. Compared to overall obesity, central obesity (abdominal fat accumulation) has a greater impact on gallstone formation, as increases in waist circumference and waist-to-hip ratio are significantly associated with gallstone prevalence[19]. Insulin resistance plays a pivotal role in gallstone formation by inhibiting bile acid synthesis, increasing cholesterol supersaturation in bile, and impairing gallbladder motility through reduced cholecystokinin (CCK) responsiveness, which leads to delayed gallbladder emptying and bile stasis, ultimately fostering an optimal environment for gallstone formation[4, 20]. Multiple studies have demonstrated a direct association between gallbladder motility dysfunction and gallstone formation[21, 22]. As epidemiological evidence on metabolic dysfunction-associated steatotic liver disease (MASLD) continues to accumulate, the interplay between metabolic dysfunction, liver fibrosis, and gallstone formation has attracted increasing attention. Research indicates that gallstone prevalence is higher among MASLD patients than non-MASLD individuals, with gallbladder contractile function progressively declining as liver fibrosis progresses[23]. Liver fibrosis alters bile acid synthesis and secretion rhythms and disrupts neural reflex regulation between the liver and gallbladder, thereby exacerbating gallstone formation[24]. Furthermore, the chronic low-grade inflammatory state in MASLD patients may modify the gallbladder microenvironment, exacerbating chronic gallbladder wall inflammation and mucin hypersecretion, which in turn promotes gallstone formation[25]. However, prior research has predominantly examined individual metabolic abnormalities or MASLD-associated gallstone risk, lacking a holistic assessment that incorporates overall metabolic burden and liver fibrosis severity. The novelty of this study lies in its pioneering evaluation of metabolic dysfunction-associated liver fibrosis, as assessed by the MAF-5 score, in relation to gallstone prevalence. Our findings establish a robust and independent positive association between MAF-5 scores and gallstone prevalence. The MAF-5 score encapsulates both metabolic burden and liver fibrosis severity, offering a more comprehensive risk assessment tool than individual metabolic markers. These findings provide novel insights into the early identification of gallstone risk in metabolically abnormal populations.

From a biological standpoint, the association between MAF-5 scores and gallstone prevalence is driven by a complex pathological network encompassing metabolic dysfunction, liver fibrosis, gallbladder motility impairment, and bile acid dysregulation. First, an elevated MAF-5 score signifies an increased metabolic burden, typified by obesity, insulin resistance, and diabetes. These metabolic disturbances inhibit CYP7A1, a key enzyme in bile acid synthesis, via insulin signaling pathways, resulting in decreased bile acid secretion and enhanced cholesterol synthesis[26, 27]. As a result, an imbalance between bile acids and cholesterol facilitates cholesterol crystal precipitation, establishing the biochemical basis for gallstone formation[28]. Moreover, liver fibrosis, as indicated by MAF-5 scores, is strongly associated with gallbladder motility dysfunction. With fibrosis progression, neural reflex pathways between the liver and gallbladder deteriorate, diminishing the gallbladder’s responsiveness to food intake stimuli and causing delayed gallbladder emptying and bile stasis, thereby fostering the kinetic environment for gallstone formation[29]. Studies have shown that gallbladder contractility is markedly reduced in MASLD patients with fibrosis compared to those with mild or no fibrosis, reinforcing the direct link between liver fibrosis and gallbladder motility impairment[30].

Chronic low-grade inflammation is a defining characteristic of individuals with elevated MAF-5 scores and plays a pivotal role in gallstone pathogenesis. Pro-inflammatory cytokines, including TNF-α and IL-6, secreted by adipose tissue, enter the liver and gallbladder through the portal circulation, inducing chronic inflammation of the gallbladder wall[31, 32]. Sustained inflammatory stimulation prompts gallbladder epithelial cells to secrete excess mucin, generating a pro-crystallization matrix that facilitates cholesterol crystal nucleation—a crucial step in gallstone formation[32, 33]. Furthermore, in diabetic individuals, diminished gallbladder responsiveness to cholecystokinin (CCK), excessive collagen deposition in the gallbladder wall, and reduced compliance exacerbate gallbladder motility dysfunction, thereby contributing to gallstone formation throughout disease progression[34]. Thus, an elevated MAF-5 score reflects a heightened metabolic burden, advanced liver fibrosis, and exacerbated chronic inflammation, reinforcing the “chemical-dynamic-inflammatory” triad as a key mechanistic driver of gallstone formation. This mechanistic framework provides a biological rationale for the linear association observed in this study.

Our findings broaden the metabolic etiology of gallstones by highlighting the contribution of metabolic dysfunction-associated liver fibrosis to gallstone formation. These results underscore the importance of early intervention in high-risk populations. Nurses play a crucial role in patient education, lifestyle modification, and postoperative care in gallstone disease management. Research suggests that nurse-led interventions focusing on dietary counseling, weight management, and physical activity promotion significantly reduce the risk of gallstone formation and recurrence[3, 35]. Additionally, perioperative nursing care is essential for patients undergoing cholecystectomy, as adequate nursing interventions can help prevent complications such as bile leaks and infections, improving patient outcomes[1]. Given the metabolic nature of gallstone disease, integrating nursing-based metabolic health education into clinical practice could enhance long-term prevention efforts.

These findings suggest that health management in metabolically abnormal populations should extend beyond traditional risk factors, such as weight control and glucose regulation, to include non-invasive liver fibrosis screening. For individuals with elevated MAF-5 scores, routine gallbladder ultrasound screening should be implemented as part of an integrated “metabolism-liver-gallbladder” strategy for early detection and intervention, ultimately aiming to prevent gallstone formation through a metabolic health-centered approach.

Nonetheless, this study has several limitations. First, the cross-sectional design precludes causal inference, underscoring the need for longitudinal cohort studies to validate these findings. Second, gallstone diagnosis was based on self-reported physician confirmation, potentially introducing recall bias; future studies should integrate imaging and biochemical assessments to enhance diagnostic accuracy. Third, key variables such as dietary patterns and family history were not accounted for in the model, leaving the potential for residual confounding. Finally, given that the study population was drawn from a U.S. cohort, further research is needed to evaluate the generalizability of these findings across diverse ethnic and geographic populations.

## 5. Conclusion

Utilizing NHANES 2017–2020 data, this study established a significant positive association between MAF-5 scores and gallstone prevalence, which persisted after adjustment for multiple confounders. These findings indicate that the MAF-5 score could function not only as a biomarker for evaluating the risk of metabolic dysfunction-associated liver fibrosis but also as a potential screening tool for identifying individuals at high prevalence of gallstones. This study offers novel insights into gallbladder health management in metabolically dysregulated populations, serving as a valuable reference for early detection and targeted prevention strategies.

## List of Abbreviations

ALT: Alanine aminotransferase
AST: Aspartate aminotransferase
BMI: Body mass index
CCK: Cholecystokinin
CI: Confidence interval
FIB-4: Fibrosis-4 Index
GPAQ: Global Physical Activity Questionnaire
HDL-C: High-density lipoprotein cholesterol
IRB: Institutional Review Board
MAF-5: Metabolic Dysfunction-Associated Fibrosis Score
MASLD: Metabolic dysfunction-associated steatotic liver disease
MET: Metabolic equivalent task
NASH: Nonalcoholic steatohepatitis
NHANES: National Health and Nutrition Examination Survey
NFS: NAFLD Fibrosis Score
OR: Odds ratio
RCS: Restricted cubic spline
SD: Standard deviation
TG: Triglycerides
TNF-α: Tumor necrosis factor-alpha
WC: Waist circumference

## Acknowledgments

We acknowledge the National Center for Health Statistics of the Centers for Disease Control and Prevention for making the National Health and Nutrition Examination Survey publicly available.

## Funding

National Natural Science Foundation of China :(grant/award numbers: 32502964);

Henan Provincial Key Scientific and Technological Projects:(grant/award numbers: 252102111021).

## Availability of data and materials

The National Health and Nutrition Examination Survey dataset is publicly available at the National Center for Health Statistics of the Center for Disease Control and Prevention (https://www.cdc.gov/nchs/nhanes/).

## Author contributions

Yujian Huang: Conceptualization, Formal analysis, Data curation, Methodology, Validation, Visualization, Writing – original draft.

Junlin Lu: Formal analysis, Data curation.

Hao Wang: Project administration, Supervision, Methodology, Writing – review & editing.

## Competing interests

The authors declare no conflict of interest.

## Consent for publication

Not applicable.

## Ethics approval and consent to participate

NHANES is conducted by the Centers for Disease Control and Prevention and the National Center for Health Statistics. The National Center for Health Statistics Research Ethics Review Committee reviewed and approved the NHANES study protocol. All participants signed a written informed consent.

## Supplemental Table Catalog

**Table S1 Sensitivity Analysis of the Association Between MAF-5 Score and Gallstone Prevalence**

## Notes

### Competing Interest Statement

The authors have declared no competing interest.

